# Complex trait associations in rare diseases and impacts on Mendelian variant interpretation

**DOI:** 10.1101/2024.01.10.24301111

**Authors:** Craig Smail, Bing Ge, Marissa R. Keever-Keigher, Carl Schwendinger-Schreck, Warren Cheung, Jeffrey J. Johnston, Cassandra Barrett, Genomic Answers for Kids Consortium, Keith Feldman, Ana S.A. Cohen, Emily G. Farrow, Isabelle Thiffault, Elin Grundberg, Tomi Pastinen

**Affiliations:** Genomic Medicine Center, Department of Pediatrics, Children’s Mercy Kansas City, Kansas City, MO, USA; UKMC School of Medicine, University of Missouri Kansas City, Kansas City, MO, USA; Department of Human Genetics, McGill University, Montreal, Canada; Health Outcomes and Health Services Research, Department of Pediatrics, Children’s Mercy Kansas City, Kansas City, MO, USA; Department of Pathology and Laboratory Medicine, Children’s Mercy Kansas City, Kansas City, MO, USA; Department of Pediatrics, Children’s Mercy Kansas City, Kansas City, MO, USA

## Abstract

Emerging evidence implicates common genetic variation – aggregated into polygenic scores (PGS) – impacting the onset and phenotypic presentation of rare diseases. In this study, we quantified individual polygenic liability for 1,151 previously published PGS in a cohort of 2,374 probands enrolled in the Genomic Answers for Kids (GA4K) rare disease study, revealing widespread associations between rare disease phenotypes and PGSs for common complex diseases and traits, blood protein levels, and brain and other organ morphological measurements. We observed increased polygenic burden in probands with variants of unknown significance (VUS) compared to unaffected carrier parents. We further observed an enrichment in overlap between diagnostic and candidate rare disease genes and large-effect PGS genes. Overall, our study supports and expands on previous findings of complex trait associations in rare disease phenotypes and provides a framework for identifying novel candidate rare disease genes and in understanding variable penetrance of candidate Mendelian disease variants.

## Introduction

The Genomic Answers for Kids (GA4K) study at Children’s Mercy Research Institute is a large-scale, phenotypically diverse pediatric rare disease cohort comprising patient cases referred from 22 different hospital specialties with almost 14,000 individuals enrolled from 6,000 families currently^1,2^. Comprehensive clinical genome assessment utilizing structured phenotypes and prioritized variants from whole-exome or whole-genome sequencing are further combined with additional omic approaches with the goal of improving understanding of the genetics of rare diseases.

One such strategy is the integration of polygenic scores (PGS) to assess the contribution of common genetic variants to rare disease phenotypes. PGS approaches have been previously applied to estimate disease risk attributed to inherited common variant polygenic background – such as in severe neurodevelopmental disorders^3^; and in integrating trait-matched PGS to understand differences in disease penetrance among carriers of monogenic risk variants^4–7^. Further, differences in individual PGS liability can help resolve variable expressivity of complex, multi-phenotype rare disorders, such as risk for schizophrenia among carriers of the 22q11.2 deletion^8^.

PGS effect estimates have also highlighted a sharing of underlying causal genes in monogenic and matched common diseases and other correlated complex traits^9,10^, providing opportunities for expanded discovery of large-effect variants underlying rare disease patient cases. Integrating common variant PGSs derived from population-scale resources available from PGS Catalog^11^, here we systematically mapped the impacts of >1000 PGSs for common complex diseases, laboratory tests, organ morphological, and anthropometric traits in >500 groupings of probands defined by their rare disease phenotypes.

## Results

To map associations between rare disease phenotypes and common complex diseases, traits, and measurements we first generated a filtered set of PGS obtained from PGS Catalog (Methods; N PGS = 1,151). We calculated individual scores for each PGS in a subset of 2,374 probands enrolled in GA4K filtered for European ancestry to reflect the demographic background of the majority of individuals currently comprising PGS training cohorts (**Methods**; **Table 1**; **Figure S1**). To calculate individual PGS scores in GA4K, genotyping array data was first combined with imputation (**Methods**). Correlation of PGS variants also available in the GA4K imputed genotype callset was high (*r* = 0.99; *P* < 1×10^−16^; **Figure S2**).

**Table 1.** Summary of GA4K probands in study cohort.

| **Table 1.** Summary of GA4K probands in study cohort |  |
| --- | --- |
| Probands, N | 2,374 |
| Sex (Female), N (%) | 1,095 (46.1) |
| Age in years at enrollment, mean (SD) | 8.5 (5.6) |
| Number of HPO terms, mean (SD) | 6.5 (4.6) |
| Diagnostic status<sup>1</sup>, N (%): |  |
| Diagnostic | 629 (48.1) |
| VUS/GUS | 369 (28.2) |
| Negative | 178 (13.6) |
| Other<sup>2</sup> | 131 (10.1) |
| <sup>1.</sup> Diagnostic status for 55.4% of study cohort who have completed clinical genetic testing.<br><sup>2.</sup> Other category includes (N): partial diagnosis (86); partial genotype (45). |  |

We quantified the contribution of each of the 1,151 PGS in 574 rare disease phenotype (HPO) case/controls cohorts (N pairwise comparisons = 660,674; HPO median case N = 10). For each of these cohorts, we constructed a logistic regression model comprising PGS, sex, and first five principal components of ancestry, and further compared observed results with null distributions from permutation testing (N permutations = 10,000), yielding an empirical P-value for each HPO-PGS pair (**Methods**). From this approach we identified 824 significant HPO-PGS pairs (FDR 20%) comprising 519 PGS and 138 HPO cohorts (**Table S1**).

Categorizing HPO and PGS into tissue- and/or physiology-specific measurements and disorders, we observed the greatest number of associations for nervous system disorder HPOs with brain volume PGSs (N = 62 HPO-PGS pairs) (for example, “severe global developmental delay” (HP:0011344) and “mean isotropic volume fraction in right superior corona radiata” PGS (PGS001506, *P* = 5×10^−05^), as well as growth disorder HPOs associated with body measurement PGSs (N = 61 HPO-PGS pairs) (for example, “obesity” (HP:0001513) and “body mass index” PGS (PGS000027), *P* = 6×10^−11^) (**Figure 1**). We further observed blood protein PGSs associated broadly with multiple HPO categories (35 unique blood protein PGS across 13 of the 20 unique HPO categories). Overall, we observed at least one significant (FDR 20%) PGS association for 1,972 of 2,374 probands in the study cohort (83%). Median regression model fit (Nagelkerke’s pseudo-R^2^) was 0.78% (range = 0.4 - 4.1%) (**Figure S3**). At more stringent FDR thresholds, we observed 331 (FDR 10%) and 182 (FDR 5%) significant HPO-PGS pairs. Summary statistics for all tested HPO-PGS pairs are provided in **Table S2**.

**Figure 1.**
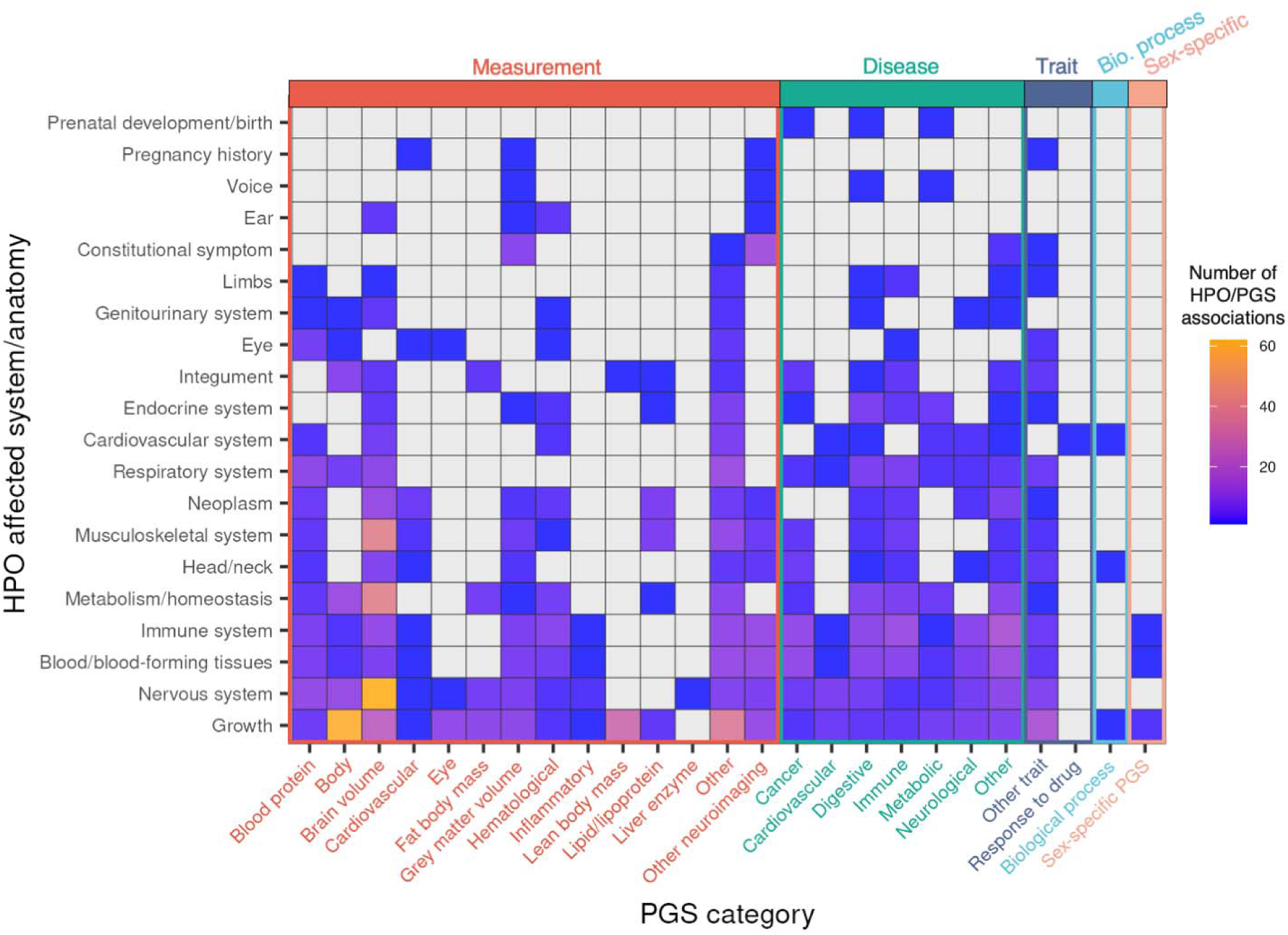
Overview of significant associations across categories of HPOs and PGSs. HPOs (y-axis) were collapsed in to corresponding affected system/anatomy. PGS terms (x-axis) were categorized using ontology metadata available from PGS Catalog and further colored by measurement (red), disease (green), other trait (blue), biological process (cyan), or sex-specific PGS (orange). Color gradient indicates number of significant associations within each HPO/PGS category pair. Gray indicates no significant associations for indicated HPO-PGS pair.

We next assessed the impact of PGS on a proband’s clinical diagnostic status. We used a logistic regression model to quantify the contribution of an increasing burden of rare disease phenotypes (HPOs) linked to PGS to the likelihood of having a clinical diagnostic status including “diagnostic”, “VUS/GUS”, “negative” classification. We repeated this analysis for increasingly stringent proband PGS Z-score thresholds. For each additional rare disease phenotype (HPO) significantly associated with a PGS (FDR 20%), we observed a modest decrease in the likelihood of “diagnostic” status (odds ratio = 0.98 [CI 0.96-0.99], *P* = 0.03) and an increase in the likelihood of “VUS/GUS” status (odds ratio = 1.02 [CI 1.01 – 1.04], *P* = 0.03) (**Figure 2**). For the most stringent PGS Z-score threshold, this effect was stronger (“diagnostic”: odds ratio = 0.87 [CI 0.81-0.94], *P* = 2×10^−04^; “VUS/GUS”: odds ratio = 1.11 [CI 1.03-1.18], *P* = 3×10^−03^). No differences were observed for “negative” cases across any PGS Z-score threshold.

We next asked whether observed results could be explained by potential challenges with clinical diagnosis of more complex patient cases (that is, a relatively larger number of rare disease phenotypes irrespective of any PGS associations). Again using a logistic regression model, we quantified the likelihood of each clinical diagnostic status associated with a count of all HPOs per proband and observed no significant associations.

**Figure 2.**
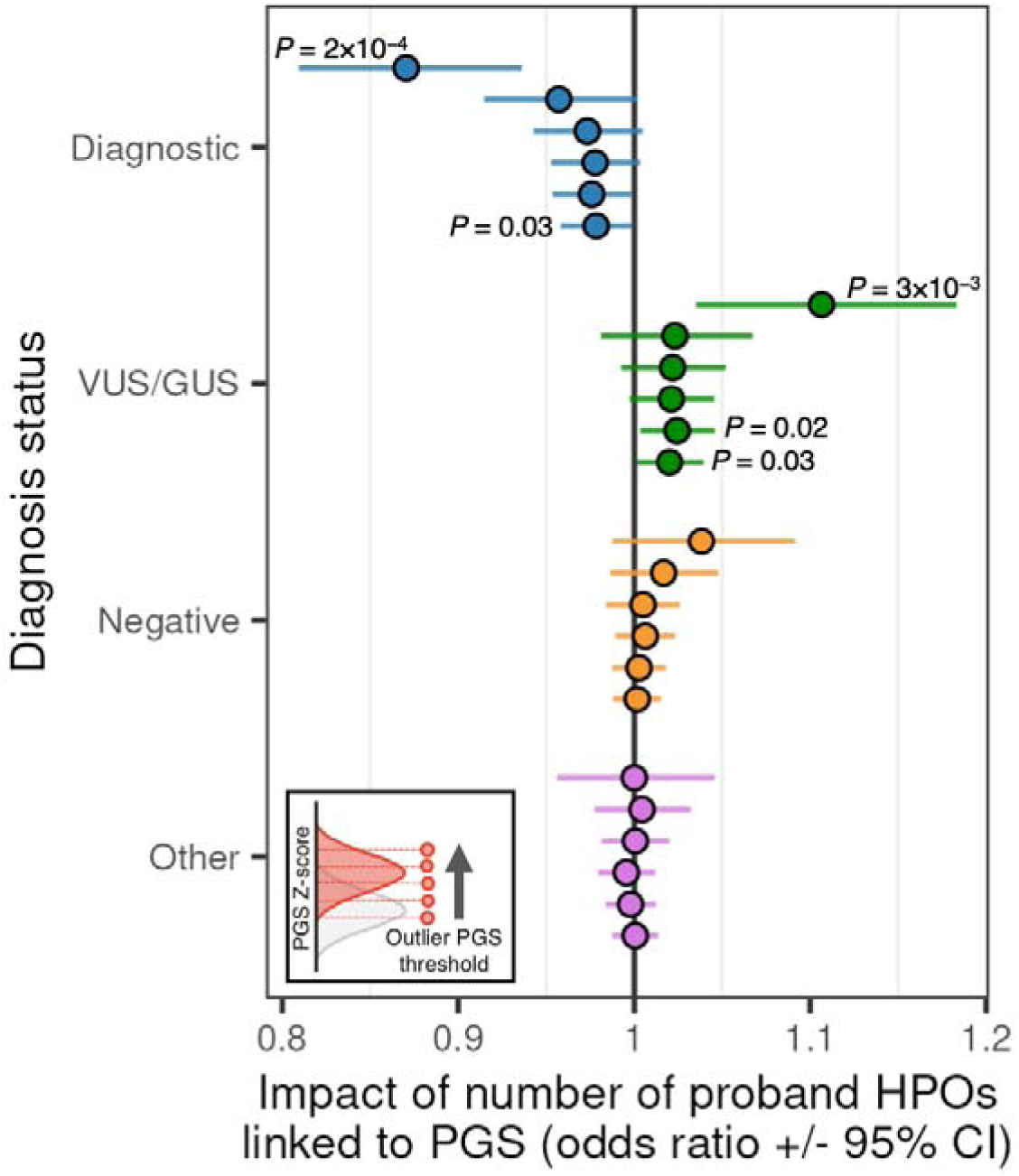
Change in likelihood of indicated clinical diagnostic status as a function of PGS-associated HPO phenotype burden. Results are displayed across more stringent PGS Z-score thresholds (from bottom to top of each diagnostic category). P-values passing a threshold of P <= 0.05 are indicated in figure.

Given previous reports showing modification of monogenic disease penetrance and severity associated with trait-relevant polygenic liability^4,6^, we hypothesized that significantly associated PGS in probands with a candidate – but presently non-diagnostic – inherited variant in a known rare disease gene (variant of unknown significance (VUS)) would exhibit increased polygenic liability compared with the PGS of their unaffected carrier parent (**Figure 3A**). In a subset of probands with an inherited, autosomal dominant (with partial penetrance) VUS and with significantly associated PGS available for the full trio (**Methods**), we calculated the difference in standard deviation (Z-score) in proband and parent PGS whereby values greater than zero indicates greater liability in probands, observing a median difference between probands and carrier parents of 0.52 SD (*P* = 3×10^-09^, Wilcoxon test against PGS permuted background), which increased at more stringent HPO-PGS significance (FDR) thresholds (FDR 10%: 0.79 SD, *P* = 7×10^-05^; FDR 5%: 0.93 SD, *P* = 4×10^-08^) (**Figure 3B**). We repeated this analysis for unaffected carrier sibling(s) where available, observing similar increased PGS liability for probands (0.65 SD, *P* = 6×10^-04^; **Figure S4A**). We observed smaller median differences in PGS when comparing probands with non-carrier parents (FDR 20%: 0.30 SD, *P* = 1×10^-06^; FDR 10%: 0.02 SD, *P* = NS; FDR 5%: -0.07 SD, *P* = NS) (**Figure 3B**).

**Figure 3.**
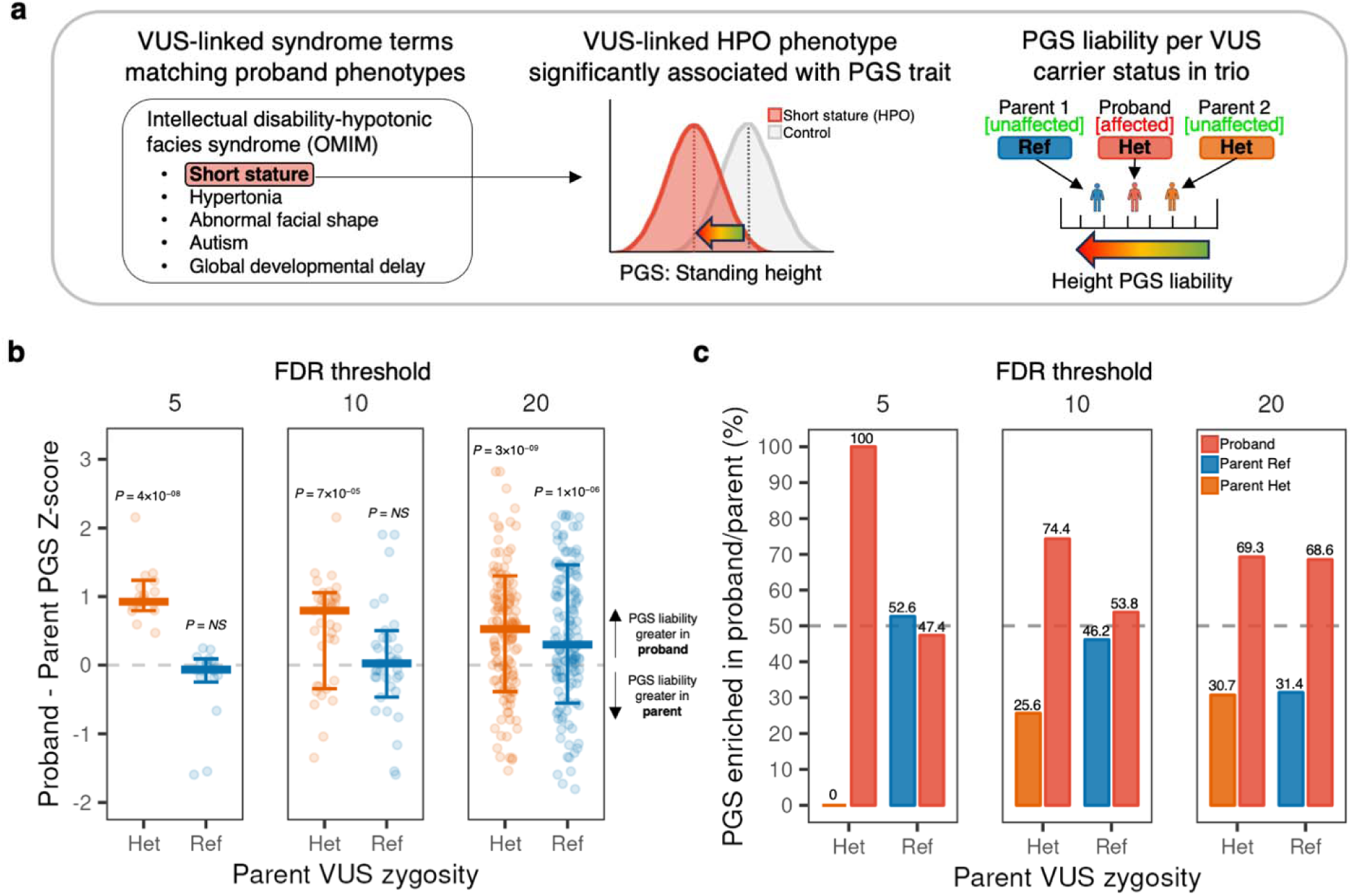
**A**. Proposed model of variable rare disease penetrance for inherited candidate pathogenic variant (VUS) in proband and unaffected carrier parent as a function of polygenic liability for an associated complex trait PGS. **B.** Proband – parent PGS Z-score/standard deviation for significantly associated PGS in probands with a clinical variant of unknown significance (VUS) compared to unaffected carrier (Het) parent (orange) and non-carrier (Ref) parent (blue). Results are stratified across HPO-PGS significance (FDR) thresholds (from right to left: FDR 20%, 10%, 5%). Points above zero indicate PGS liability is greater in the proband, whereas points below zero indicate PGS liability is greater in the indicated Het or Ref parent. Pairwise differences for PGS with direction of effect < 0 (e.g. short stature HPO and Height PGS) were inverted to enable visualization in the same figure of PGS with direction of effect > 0. **C.** Summarized within-family comparison of PGS enrichment for VUS probands and parents. Pairwise comparisons are shown for proband (red) and Het parent (orange), and proband and Ref parent (blue). Results are stratified across HPO-PGS significance (FDR) thresholds (from right to left: FDR 20%, 10%, 5%).

We next calculated within-trio comparisons across all matched PGS to quantify the proportion of observations where the proband exhibits enriched PGS liability compared with their carrier and non-carrier parent (that is, PGS is greater/less than the indicated parent with respect to associated PGS direction of effect), repeated across HPO-PGS significance (FDR) thresholds. We observed a greater proportion of increased PGS liability for probands compared with their carrier parent (PGS enriched in proband: 69.3% (FDR 20%), 74.4% (FDR 10%), 100% (FDR 5%), but did not observe the same trend when comparing with noncarrier parent (PGS enriched in proband: 68.6% (FDR 20%), 53.8% (FDR 10%), 52.6% (FDR 5%)) (**Figure 3C**). We further observed a greater proportion of increased PGS for probands compared with unaffected carrier sibling(s) (**Figure S4B**).

Next, we obtained clinical observation data for height from the electronic medical record (EHR) at the GA4K study site to compare individual PGS with growth trajectories recorded at clinical visits. We observed moderate positive correlation comparing CDC age- and sex-adjusted height Z-scores with a significantly associated PGS for height (PGS ID = “PGS000998”; **Methods**) (*r* = 0.36, *P* < 1×10^−16^) (**Figure 4**). We then integrated rare disease phenotypes (HPO terms) to annotate “short stature” (HP:0004322), “tall stature” (HP:0000098), and control cohorts. For case probands, we observed a partial overlap in height growth chart values exceeding the CDC-recommended outlier threshold for short or tall stature (height Z-score <=2 or >=2, respectively). For the short stature cohort (N = 207), 95 (46%) passed the threshold, and for tall stature cohort (N = 32), 19 (60%) passed the corresponding threshold.

For probands with observed outlier height Z-scores we observed that their trait-relevant PGS liability tended to stratify into expected direction of effect. Specifically, for the short stature cohort with outlier height Z-score (N = 95), 74 (78%) had a PGS <=0 and 21 (22%) > 0. For tall stature (N = 19), 18 (95%) had a PGS >= 0 and 1 (5%) < 0. This finding highlights the potential for enhanced rare disease phenotyping through integrating EHR and PGS information.

**Figure 4.**
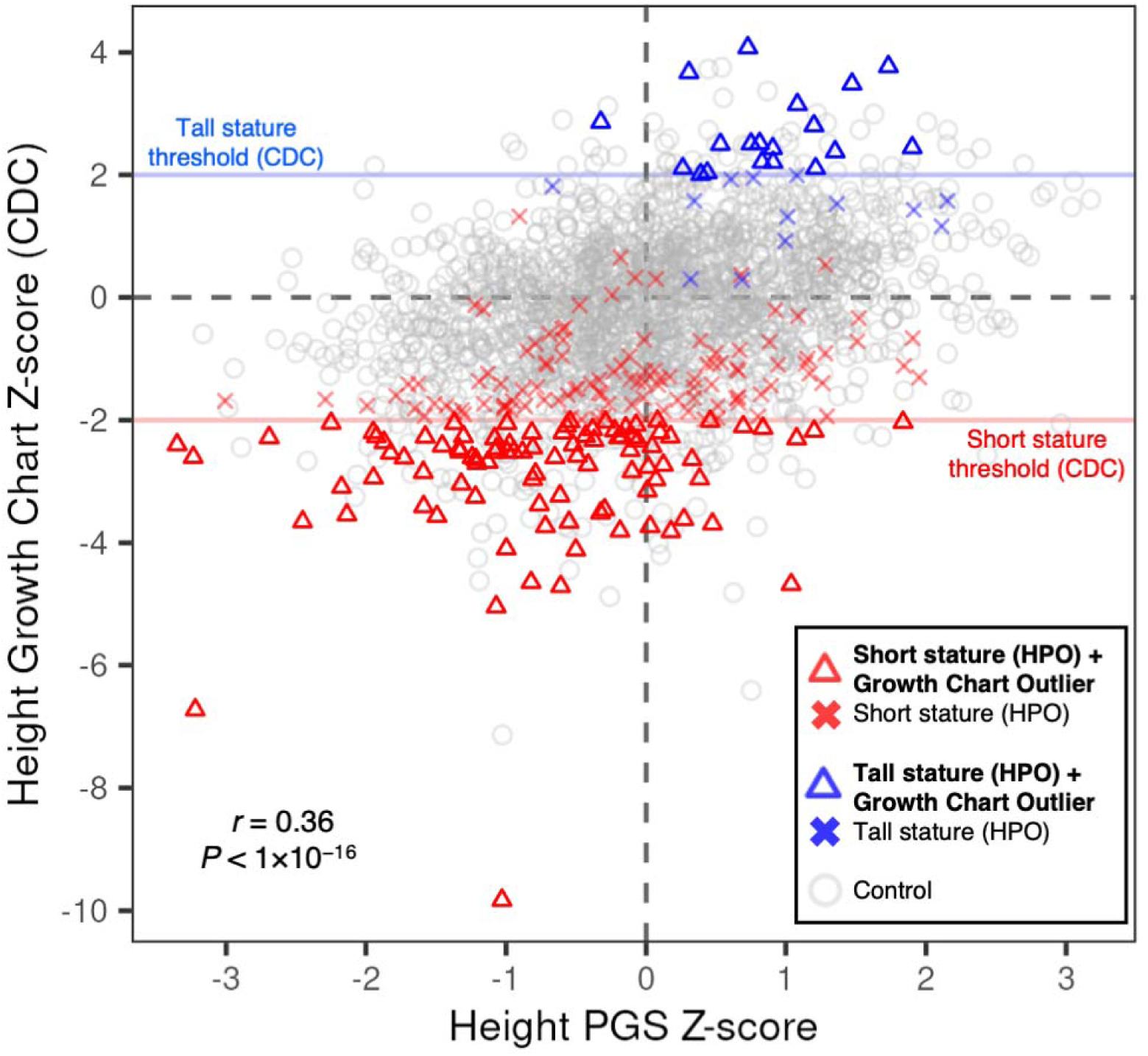
Integrating PGS and clinical growth chart data. Individual height observations indicate age and sex-adjusted Z-scores (CDC benchmarks) and are displayed on the y-axis. Individual scores for a significantly associated height PGS are displayed on the x-axis. Red horizontal line indicates short stature threshold (CDC). Blue horizontal line indicates tall stature threshold (CDC). Probands with a short stature HPO in GA4K are indicated by a red triangle if observed height value passes CDC threshold for short stature, or a red cross if not passing this threshold. Probands with tall stature HPO are indicated by a blue triangle if observed height value passes CDC threshold for tall stature, or a blue cross if not passing this threshold. Gray indicates control (no short or tall stature HPO terms present in GA4K). Pearson correlation coefficient and P-value is indicated in figure.

We next assessed the overlap of clinical diagnostic or candidate (VUS) rare disease variants in genes that are also present in significantly associated PGS. Across all PGS genes previously linked to the associated HPO rare disease phenotype in OMIM or Orphanet (HPO gene associations; see **Data Availability**), we observed a 6-fold increase in cases having a diagnostic or candidate rare disease variant in a PGS gene compared to controls (Fisher’s exact test, odds ratio = 5.99 [CI 5.67–6.33], *P* < 1×10^-16^) (**Figure 5A**). We further implemented a method for ranking PGS genes to define a set of core/key genes where a rare variant has the potential to exert a relatively larger effect on disease risk, as postulated in the omnigenic model of complex traits^12^. Previous studies have shown that GWAS effect sizes tend to be larger for genes overlapping known disease-matched monogenic disorder genes^10^; given this finding, we ranked genes in each PGS using summarized effect weights for variants in a PGS that are within or proximal (+/-10 Kb) to any protein coding gene (**Methods**). We observed for case probands an increasing enrichment in overlap in clinical diagnostic or candidate variants in genes with higher rank in the PGS. For example, for the top 0.01% effect rank of PGS genes we observed a 20-fold increase in clinical diagnostic or candidate variant overlap in cases compared to controls (Fisher’s exact test, odds ratio = 20.33 [CI 10.67–37.29], *P* = 2×10^-15^). This effect was stronger when increasing the stringency of HPO-PGS significance threshold (FDR 10%: odds ratio = 45.59 [CI 19.56–109.49], P < 1×10^-16^; FDR 5%: odds ratio = 49.85 [CI 19.78–136.06], P < 1×10^-16^) (**Figure S5**).

**Figure 5.**
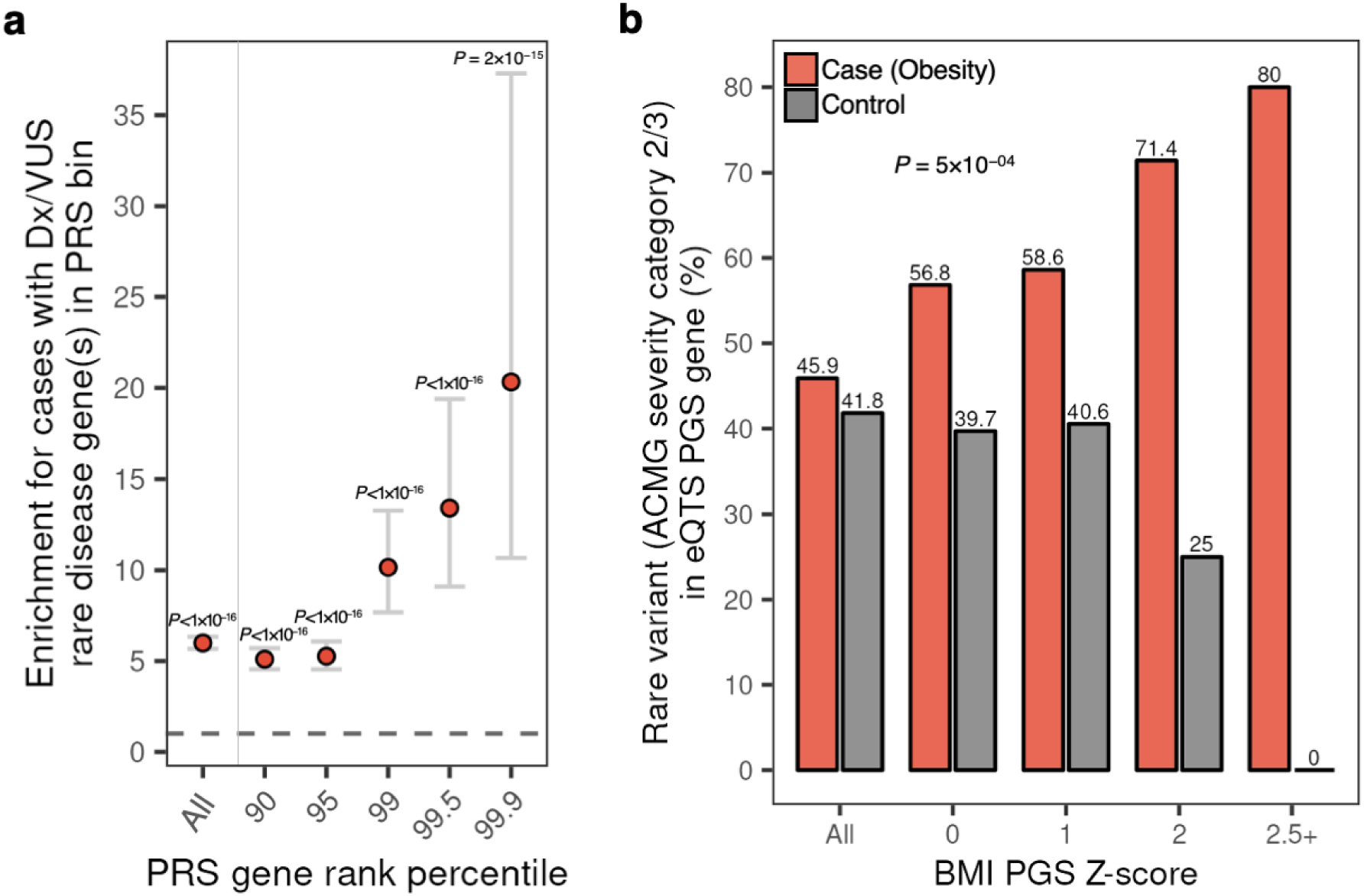
Quantifying overlap between rare disease variant annotations and putative core/key genes in associated PGS in cases and controls. **A.** Enrichment for HPO cases versus controls with a diagnostic or candidate (VUS) rare disease variant in an associated PGS gene with indicated effect rank percentile in PGS. **B.** Proportion of cases (obesity cohort) and controls with rare variants prioritized in to ACMG severity categories 2 or 3 in an obesity eQTS gene as a function of individual liability for a significantly associated polygenic score for body mass index (BMI).

Using an orthogonal metric to define putative trait-relevant core/key PGS genes, we focused on trait-matched expression quantitative trait scores (eQTS) genes (eQTLGen Consortium, see Data Availability). eQTS measures the correlation between the expression of a given gene and a polygenic score, aggregating regulatory effects on expression for both cis-and trans-acting variants^13^. The genes with the strongest correlation for a given trait are then defined as putative core/key genes. We assessed the likelihood of a proband harboring a potential large-effect rare variant (ACMG severity category 2 or 3; **Methods**) in an eQTS gene across increasingly more stringent PGS liability thresholds. Focusing on a subset of the HPO obesity case/control cohort with clinical sequencing data available (N = 1,168), we identified candidate rare variants in a set of 34 eQTS genes significantly associated with obesity or extreme BMI (**Methods**). Across the full cohort, 28 of 61 (46%) cases harbored a potential large-effect rare variant in a trait-relevant eQTS gene and 463 of 1,107 (42%) controls (*P* = 0.59).

When then considering individual polygenic liability for a significantly associated PGS trait (PGS = “body mass index”) in addition to rare variant burden, we observed an increasing enrichment in the proportion of cases with rare variants in trait-relevant eQTS genes as a function of trait-relevant polygenic liability, but a depletion in controls (logistic regression, interaction *P* = 5×10^−04^). At an outlier PGS Z-score threshold of >=2, 10 of 14 (71%) cases were found to have a potentially impactful rare variant, and 6 of 24 (25%) controls (**Figure 5B**). Increasing the PGS Z-score outlier threshold further (PGS Z-score >= 2.5), we observed 4 of 5 (80%) cases with potentially impactful rare variants in trait-relevant eQTS genes and 0 of 6 controls. We repeated this analysis using the same patient cohort across six eQTS gene sets for non-matched diseases or traits and observed no significant differences between cases and controls (**Figure S6**). Of the 34 obesity-associated eQTS genes used in this analysis, only one has been previously linked to a rare Mendelian obesity disorder in OMIM or Orphanet (HPO gene associations; see **Data Availability**). These findings suggest a potential framework for the discovery of novel rare disease genes integrating rare disease-associated PGS liability, PGS gene effect estimates, and clinical rare variant annotations.

## Discussion

Recent studies have shown that integrating PGS information can help in understanding the genetic basis of rare diseases^3,14–17^. Here, we implemented a pipeline for the systematic assessment of PGS associations in a large-scale, phenotypically diverse pediatric rare disease cohort, mapping patient phenotypes to over 500 complex trait PGSs. We demonstrated how individual polygenic liability is enriched in proband carriers of inherited, clinically prioritized variants of unknown significance (VUS) compared to unaffected carrier parents. Furthermore, using two separate metrics to define putative core/key PGS genes, we identified strong overlap between clinical diagnostic or candidate rare disease genes and those with large estimated effects in significantly associated PGS. Finally, we proposed a framework for the prioritization of candidate variants in novel rare disease genes.

The accurate classification of VUS pathogenicity is an ongoing and growing problem in the field^18^. Our findings provide evidence that integrating information from proband and parent polygenic backgrounds for associated complex traits could be a useful addition to variant annotation workflows. We focused on a set of inherited variants with high suspicion of pathogenicity resulting from expert clinical geneticist review and suspected to cause disease through autosomal dominance with partial penetrance. An explanation for why some phenotypes comprising the nominated syndrome are not observed in unaffected carrier parents could be helpful in the further annotation of these candidate diagnostic variants. For the set of inherited VUS studied here, observed differences in polygenic background between probands and unaffected carrier parents might explain variable presentation of the candidate rare disorder. Additionally, with relevance to ongoing efforts to experimentally catalog the functional impact of VUS and other possible disease variants using multiplexed assays of variant effect (MAVEs) or similar platforms^19,20^, our results suggest that the utility of these approaches might be maximized when using patient-specific cell lines to reflect potentially relevant polygenic background associated with rare disease presentation.

One key limitation of our approach is the focus on the subset of GA4K individuals with European ancestry. This constraint reflects bias toward this ancestry group in available GWAS and PGS training cohorts^11^, as well as in the GA4K study cohort itself. Future updates to our work will benefit from ongoing efforts in multi-ancestry PGS development^21^ and targeted approaches for equitable outreach and enrollment in GA4K^22^. Furthermore, our approach associates single standardized phenotype terms (HPO) with PGS, whereas our results showing a mismatch in growth disorders HPOs and height growth chart Z-scores from EHR data suggest an opportunity for improved rare disease phenotyping through integrating multiple data modalities.

Combined, our study suggests that future advances in the diagnosis of rare diseases will be enabled by considering the full frequency spectrum of genetic variation present in patient cases.

## Methods

### GA4K genotype imputation

Genotyping of the full GA4K study cohort was performed using the Avera Global Screening Array (24v1-0_A1, stranded). Genotyping array variant calls in hg38 build for individuals with genotyping call rate >= 90% were then used as input for imputation using the TOPMed imputation server with the r2 (1.0.0) reference panel^23^. From the full set of 548,291 measured genotypes, 533,837 (97.43%) overlapped with the reference set.

### GA4K ancestry inference

Principal component analysis was performed on a subset of LD and MAF pruned variants from chromosome 1 using plink (version 2)^24^ (“indep-pairwise” flag, values: 1000, 50, 0.05; “maf” flag, value: 0.01) and flashpcaR (version 2.0.1)^25^. Using a subset of known ancestry labels, the full GA4K cohort was filtered to define a European (EUR) ancestry cohort composed of individuals with known EUR ancestry and those inferred from manual inspection of a principal components plot (PC1 > 0, PC2 < 0.005).

### Filtering PGS and calculating individual scores

Individual-level PGSs for GA4K participants were calculated using PGS weights obtained from The Polygenic Score (PGS) Catalog^11^ (see **Data Availability**). From a starting set of 3,334 PGS downloaded from PGS Catalog (February 2023), PGS measuring the same trait or disease were filtered by retaining the PGS with the greatest number of variants. Further, PGS measuring lifestyle phenotypes such as diet were removed. PGS containing duplicated variants (more than one weight listed for the same variant) were removed. PGS variants were converted to hg38 coordinates using dbSNP^26^ (version 155) where necessary. 1,151 PGS remained after applying these filtering criteria. PGS were linked to category labels using the PGS ontology ID and trait category annotations available from PGS Catalog. PGS variants were restricted to autosomes only and variants mapping to the HLA region were removed. PGSs were calculated using plink (version 1.9)^24^ (“sum” flag) on all variants available in the GA4K imputed genotype callset with R^2^>=0.8 (“exclude-if-info” flag). PGS scores were converted to Z-scores within each PGS in the full GA4K EUR ancestry cohort (proband and other family members, N = 7,436). Individuals with an extreme outlier PGS Z-score (abs(PGS Z-score)>=10) in one or more PGS were removed from further analysis. This cohort was then filtered for probands only (N = 2,374). The remaining samples were retained for analysis using mother, father, and/or sibling PGS data.

### Identifying significant HPO-PGS associations

PGSs were associated with rare disease case sub-cohorts using HPO IDs for any ID with a minimum proband count 5 among the 2,374 proband study cohort (N = 574 sub-cohorts). For each HPO sub-cohort and PGS (N HPO = 574; N PRS = 1,151; N = 660,674 pairwise comparisons) a logistic regression model was used with HPO case/control status as the response variable and PGS Z-score, first five principal components of ancestry (see “GA4K inferred ancestry”), and sex as predictor variables. Model fit was assessed with Nagelkerke’s R squared using the NagelkerkeR2 function in the fmsb package (version 0.7.5) in R. An empirical P-value was calculated for each pairwise comparison using a simulated null distribution of logistic regression abs(Z-statistics) across 10,000 permutations of random case/control label reassignment (retaining relative HPO case/control counts) within each HPO-PGS pair (approximately 6.6B total model calculations). Predictor variables remained as described above. Empirical P-values were generated in R using the empPvals function available in the qvalue package (version 2.30.0)^27^. The empirical P-value is defined here as the fraction of null logistic regression abs(Z-statistics) that match or exceed the observed abs(Z-statistic) (plus an integer constant) for a given HPO-PGS pair. In this way, the minimum possible empirical P-value for a given HPO-PGS pair in the present analysis is 1×10^-04^ (i.e., the observed statistic is not matched or exceeded by any of the 10,000 null statistics from permutations). The expectation of this procedure is that the distribution of null empirical P-values follows a uniform distribution^28^; however, when combining empirical P-values across all HPO-PGS pairs it was observed that this distribution was not uniform (Kolmogorov-Smirnov test) and that the estimate of null statistics was affected by the relative sample size of each HPO sub-cohort. Therefore, the false discovery rate (FDR) was calculated within each HPO case sub-cohort separately (N = 574 cohorts) to conform with the expected uniform distribution of empirical P-values.

### Linking PGS variants to genes and effect size ranking

PGS variants within or proximal (+/-10 Kb) to any protein coding gene were identified using bedtools (version 2.29.1)^29^ (“window” flag; value: 10,000). Gene coordinates and annotations were downloaded from GENCODE (file: gencode.v26.GRCh38.genes.gtf) (see **Data Availability).** For each PGS, genes were ranked by effect size by selecting the linked variant (i.e. within the gene body or +/-10 Kb) with the maximum absolute PGS effect weight.

### GA4K clinical sequencing and phenotypic data

Clinical whole-exome (WES) or whole-genome (WGS) variant calls, standardized phenotype codes (Human Phenotype Ontology (HPO)^30^), and diagnostic status were obtained from the GA4K study repository as previously described^1^. HPO terms were summarized for visualization where indicated using ontology parent terms specified in HPO (see **Data Availability**), extracted using the get_ancestors function available in the ontologyIndex package (version 2.11)^31^ in R.

For the analysis of probands with clinical diagnostic status of variant of unknown significance (VUS), probands were selected if their PGS-associated HPO(s) matched any of the disease phenotypes for candidate syndromes returned by clinical annotation performed by clinical geneticists. This annotation pipeline prioritizes disease variants and summarizes matching phenotypes by aggregating across multiple knowledge bases such as OMIM, ACMG, and ClinVar. VUS cases were restricted to single inherited variants (i.e. VUS is present in one parent), autosomal only, annotated as autosomal dominant with partial penetrance, and where associated PGSs were available for the full trio. Compound heterozygous VUS events were removed. Parent VUS zygosity was obtained from clinical WES/WGS available in the GA4K study repository. Carrier parents were confirmed to be unaffected for the matching VUS phenotype(s) by clinical geneticist chart review.

Height and weight observations, including measure date/time and patient ancestry, were obtained from the electronic medical record at the GA4K study site at Children’s Mercy Hospital. Observations were filtered for “White” ancestry label. The R package cdcanthro (version 0.1.1) was used to calculate age and sex adjusted Z-scores for height and weight using CDC guidelines. A larger patient cohort (N = 3,073) was used in order to maximize the number of age-matched observations available and therefore the accuracy of growth Z-scores. Further, BMI Z-scores were calculated using the formula Weight/(Height/100)^2. The average (median) Z-score for each measurement was computed for individuals with more than one observation time point.

### Assessing rare variant burden in large-effect PGS genes

To assess the landscape of rare variants in putative core/key PGS genes (eQTS genes) for obesity, rare variant impact annotations were obtained using Variant Effect Predictor (VEP) (version 98_38)^32^ and allele frequencies from gnomAD (version 3)^33^. Rare variants were defined as those with gnomAD MAF < 1% (and including variants absent in gnomAD) and were further categorized using ACMG variant severity guidelines^34^. Variants annotated in category 2 or 3 were retained. Rare variants in category 2 are defined as: nonsense; disruption of stop; loss of initiation; splice junction; donor/acceptor (AG/GT); frameshift; whole transcript deletion. Category 3 variants are defined as: missense; in-frame in/del including whole exon; intronic or synonymous variant possibly affecting splicing (in polypyrimidine tract, five_prime_exonic, five_prime_flank, three_prime_exonic, five_prime_intronic, three_prime_flank); any variant in a mitochondrial gene.

eQTS genes (FDR < 5%) were obtained from eQTLGen Consortium (see **Data Availability**), and subset using the trait column for the terms “obesity” or “extreme_bmi”. To create a set of non-relevant traits, the trait column was subset for the terms “asthma”, “celiac disease”, “juvenile idiopathic arthritis”, “primary biliary cirrhosis”, “educational attainment”, or “coronary artery disease”. In this analysis, the case cohort was defined as those with a HPO term for “obesity” (HP:0001513), “increased body weight” (HP:0004324), and/or “overweight” (HP:0025502), as well as having a BMI growth chart Z-score > 2. Controls were defined as any sample without any of these HPO terms, as well as having a BMI growth chart Z-score of < 1.

All statistical analyses were performed in R (version 4.2.1). Figures were generated using ggplot2 (version 3.4.0)^35^.

## Data Availability

GA4K data is available via dbGAP using accession number phs002206.v4.p1 and AnVIL at https://anvilproject.org/data/studies/phs002206. Polygenic scores are available from PGS Catalog at https://www.pgscatalog.org/. Gene annotations are available from GENCODE (version 26) at https://www.gencodegenes.org/human/release_26.html. eQTS genes are available from eQTLGen Consortium at https://www.eqtlgen.org/eqts.html. Human Phenotype Ontology (HPO) in OBO format is available at https://hpo.jax.org/app/data/ontology. HPO phenotype-to-gene annotations are available at https://hpo.jax.org/app/data/annotations.

## Ethical declarations

The study was approved by the Children’s Mercy Institutional Review Board (IRB) (Study # 11120514). Informed written consent was obtained from all participants prior to study inclusion.

## Acknowledgements

C.S. is supported by NIH grants R35GM146966 and R21HG012422. The work of GA4K was made possible in part by generous gifts to Children’s Mercy Research Institute at Children’s Mercy Kansas City.

