## Supplemental Figures for "Complex trait associations in rare diseases and impacts on Mendelian variant interpretation"

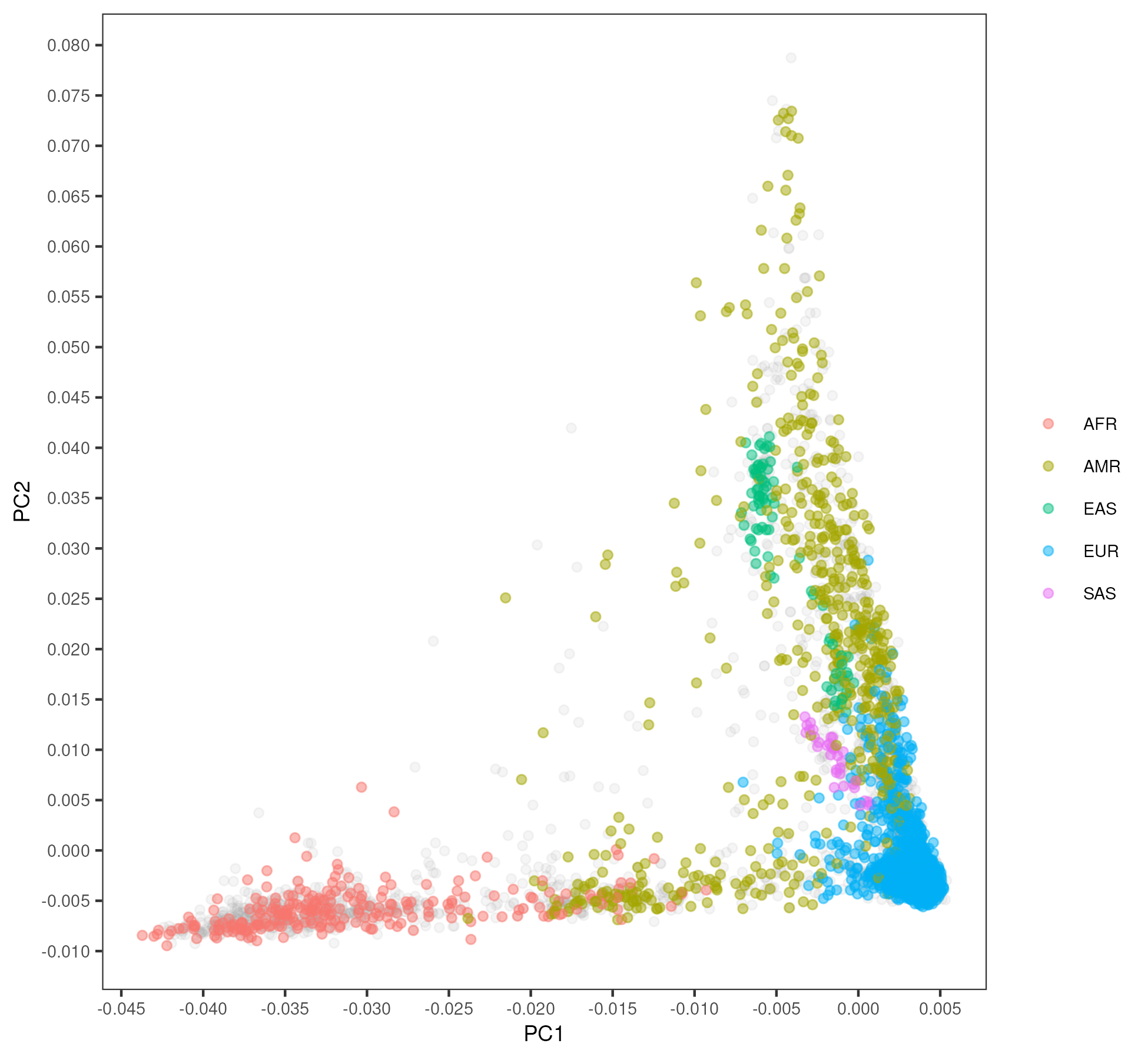

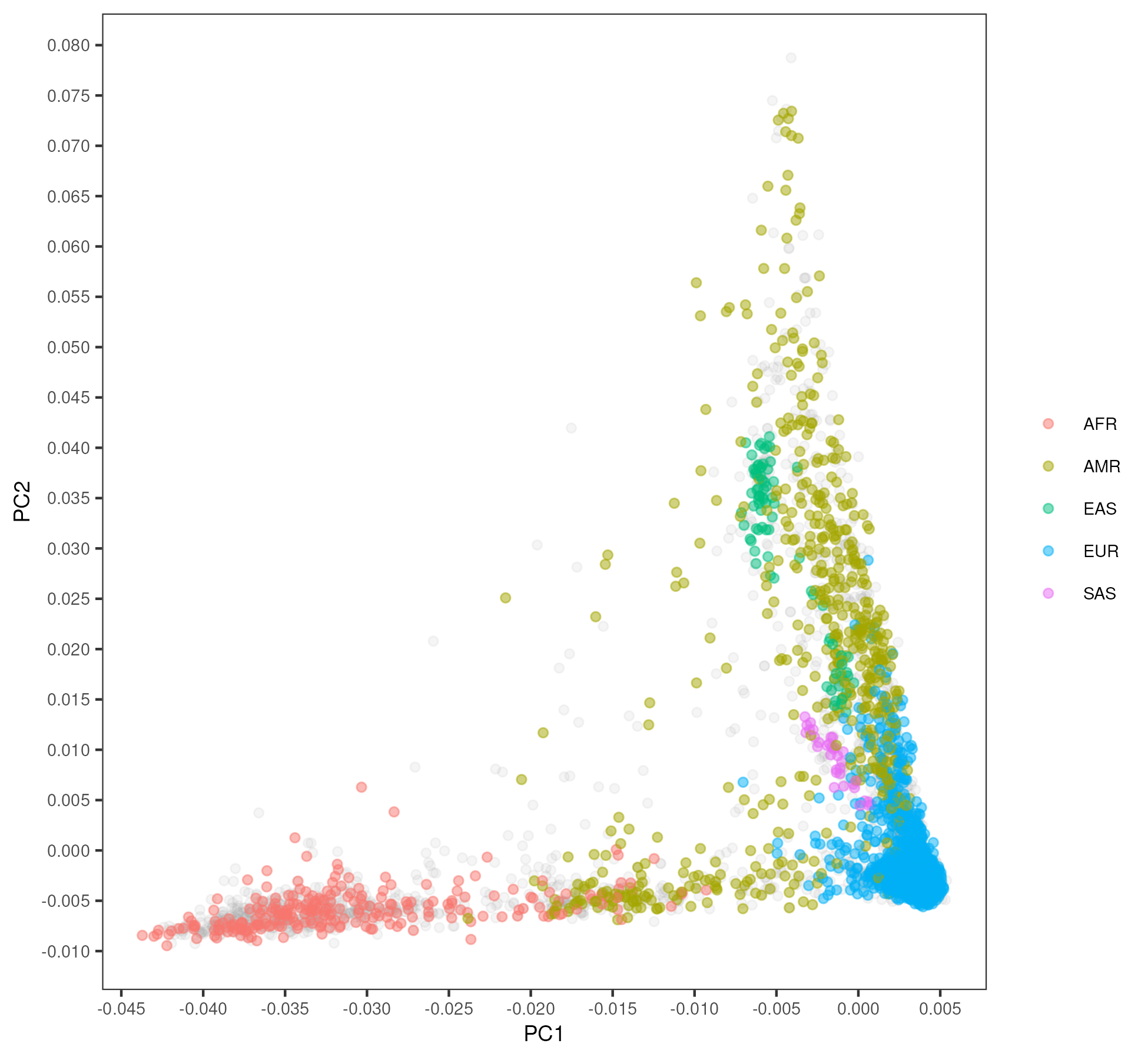

**Supplementary Figure 1.** Principal components analysis in full GA4K cohort. Dots with color indicate known ancestry labels.

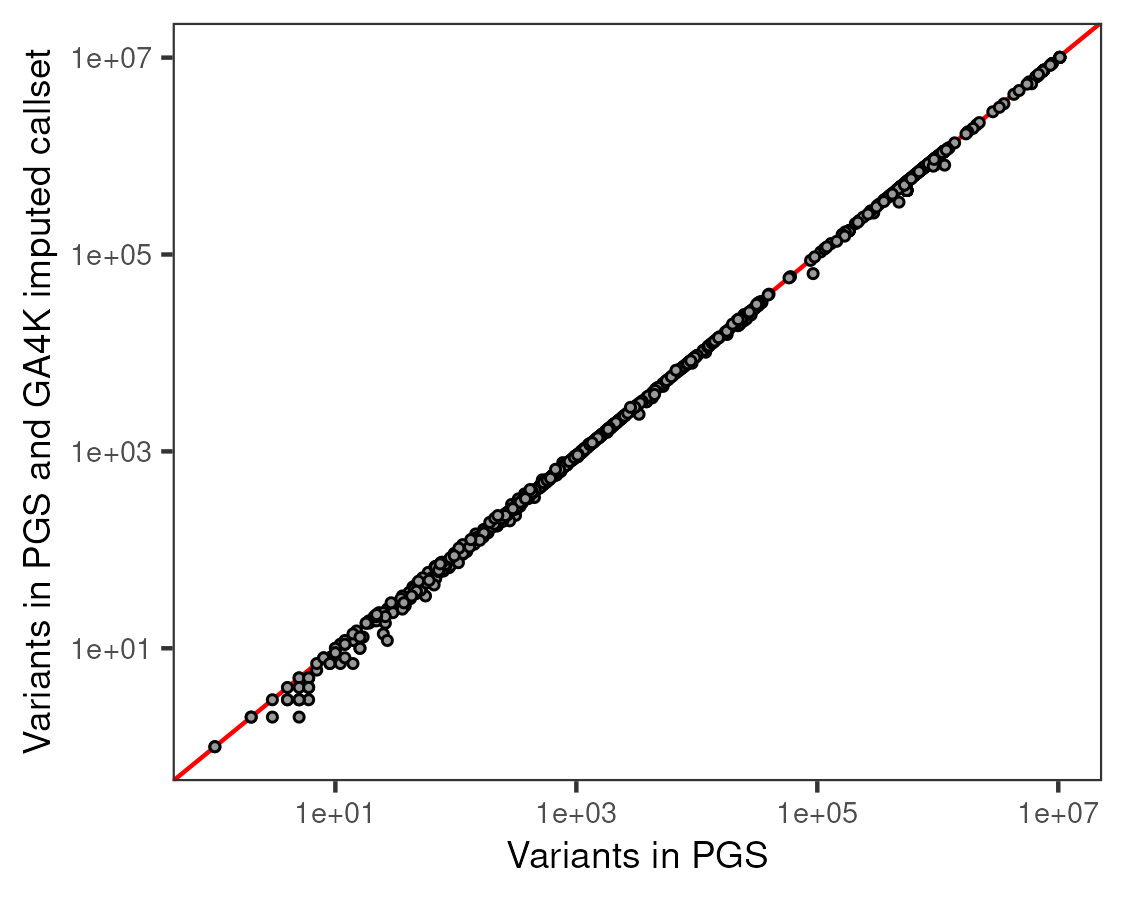

*r =* 0.99

*P < 1×10^-16^*

**Supplementary Figure 2.** Correlation between number of variants in each PGS (x-axis) and mean number of non-missing PGS variants in the GA4K imputed callset (filtered for study probands only) for each corresponding PGS (y-axis).

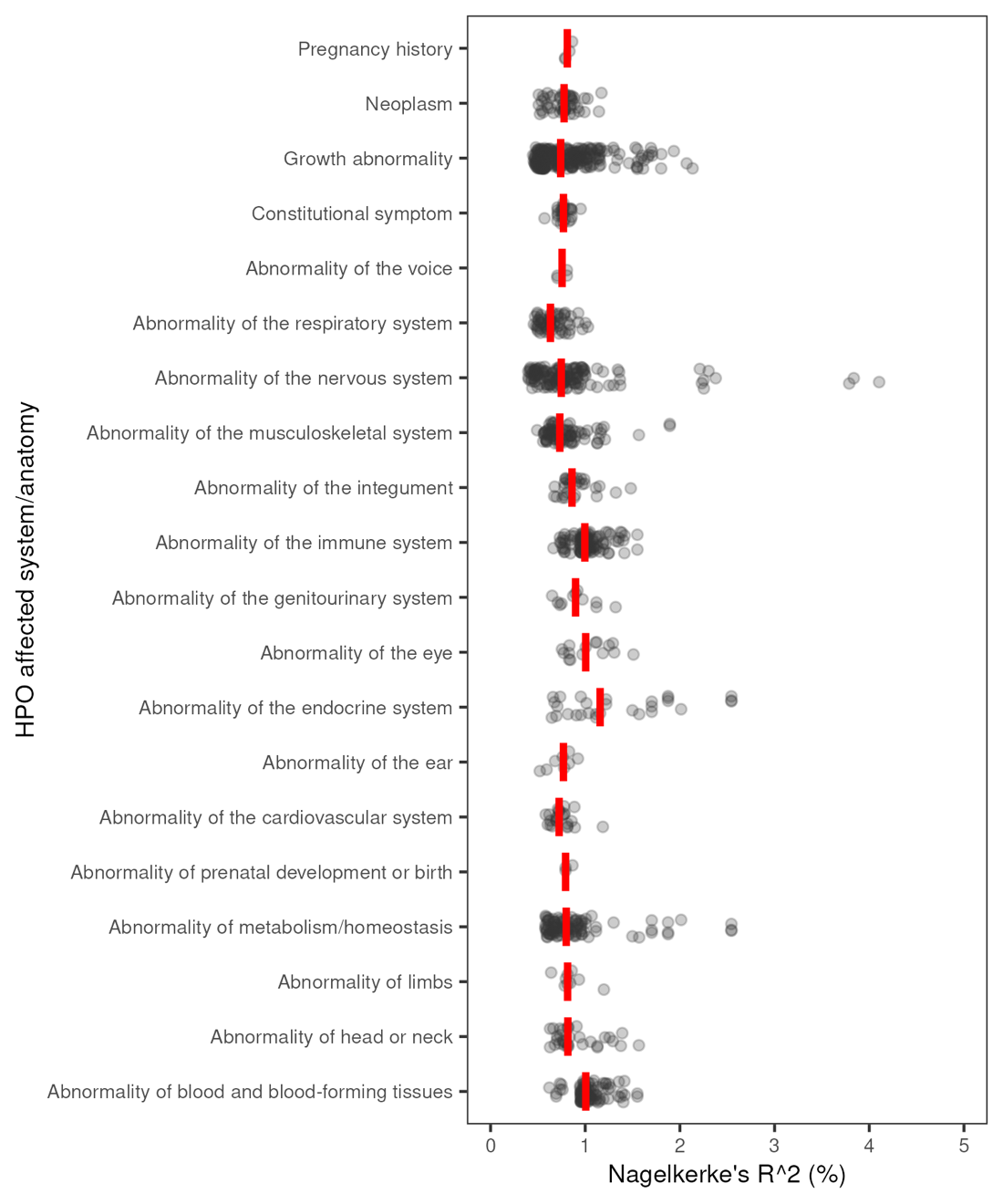

**Supplementary Figure 3.** Overview of logistic regression model fit (Nagelkerke's R squared) for significant HPO/PGS pairs, summarized by parent HPO category. Red lines indicate median.

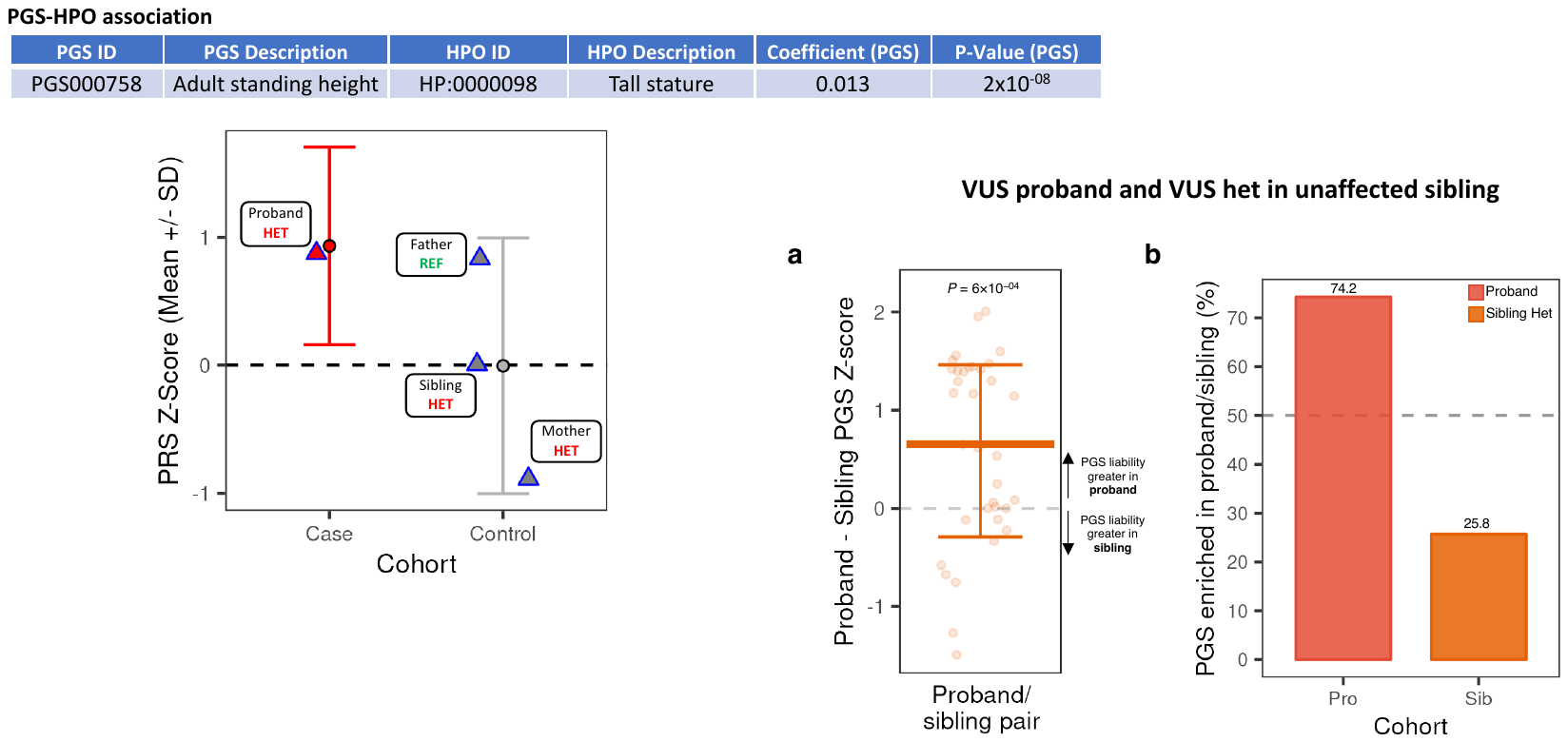

**Supplementary Figure 4.** PGS for selected probands with a variant of unknown significant and unaffected carrier sibling(s). **A.** Proband – sibling PGS Z-score/standard deviation for significantly associated PGS in probands with a clinical variant of unknown significance (VUS) compared to unaffected carrier sibling(s). Points above zero indicate PGS liability is greater in the proband, whereas points below zero indicate PGS liability is greater in sibling. Pairwise differences for PGS with direction of effect < 0 (e.g. short stature HPO and Height PGS) were inverted to enable visualization in the same figure of PGS with direction of effect > 0. **B.** Within-family (proband compared with sibling(s)) comparison of PGS enrichment for each significantly associated PGS.

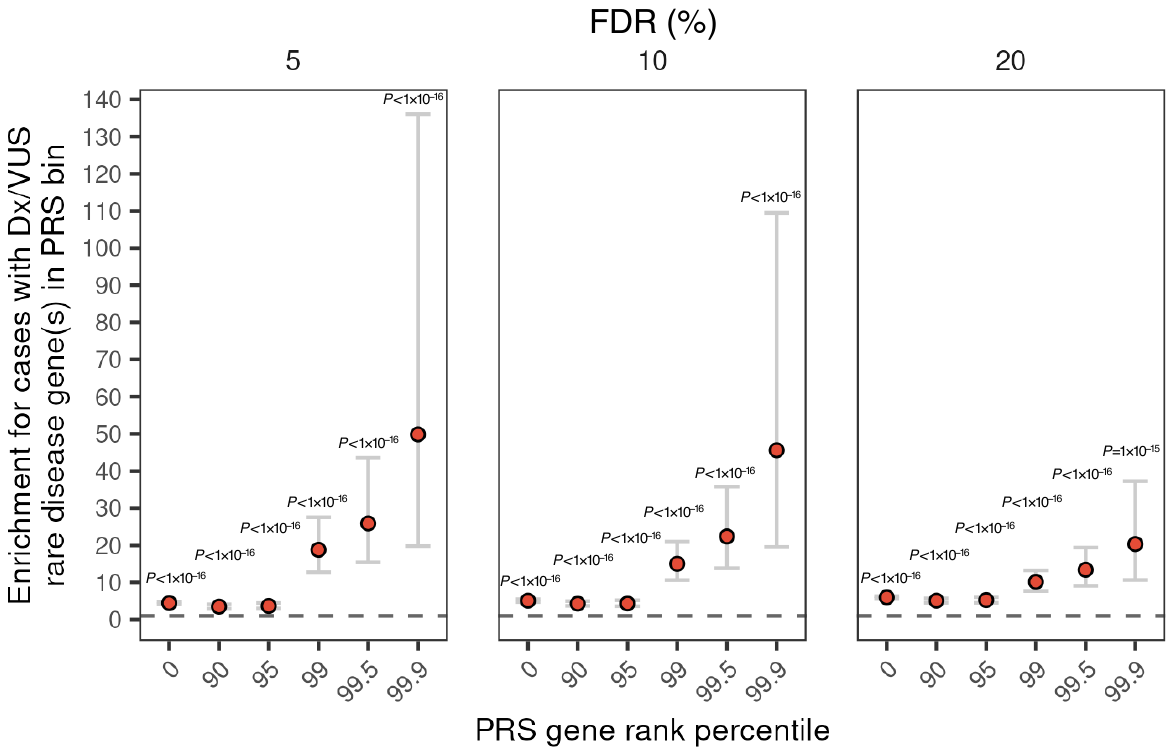

**Supplementary Figure 5.** Enrichment for HPO cases versus controls with a diagnostic or candidate (VUS) rare disease variant in an associated PGS gene with indicated effect rank percentile in PGS across significance (FDR) PGS-HPO thresholds.

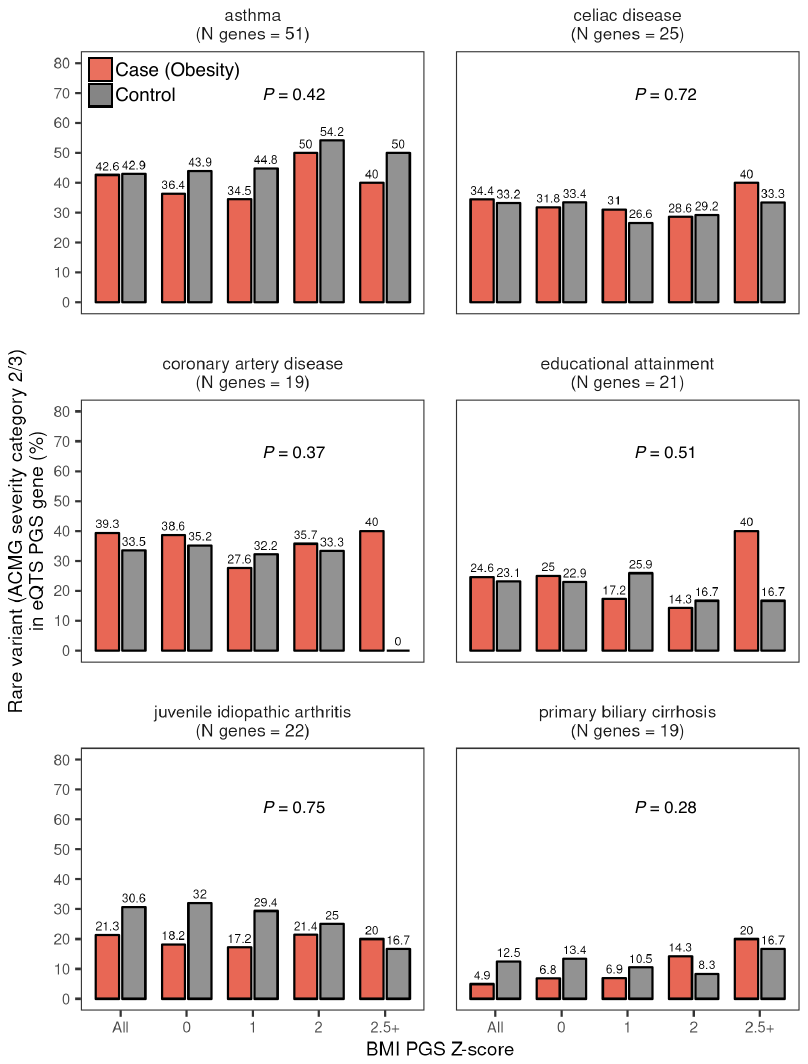

**Supplementary Figure 6.** Overlap in non-disease matched putative core/key PGS genes (eQTS genes) for cases compared to controls for rare variants prioritized in to ACMG severity categories 2 or 3.

**Genomic Answers for Kids (GA4K) Consortium Author List**

Ana S.A. Cohen^1,2,3^

Emily G. Farrow^1,3,4^

Ahmed T. Abdelmoity^4^

Joseph T. Alaimo^2,3^

Shivarajan M. Amudhavalli^3,5^

John T. Anderson^6^

Lalit Bansal^4^

Lauren Bartik^3,5^

Primo Baybayan^7^

Bradley Belden^1^

Courtney D. Berrios^1^

Rebecca L. Biswell^1^

Pawel Buczkowicz^8^

Orion Buske^8^

Shreyasee Chakraborty^7^

Warren A. Cheung^1^

Keith A. Coffman^4^

Ashley M. Cooper^4^

Laura A. Cross5

Tom Curran^9^

Thuy Tien T. Dang^4^

Mary M. Elfrink^1^

Kendra L. Engleman^5^

Erin D. Fecske^4^

Cynthia Fieser^4^

Keely Fitzgerald^4^

Emily A. Fleming^5^

Randi N. Gadea^5^

Jennifer L. Gannon^5^

Rose N. Gelineau-Morel^3,4^

Margaret Gibson^1^

Jeffrey Goldstein^4^

Elin Grundberg^1^

Kelsee Halpin^3,4^

Brian S. Harvey^6^

Bryce A. Heese^5^

Wendy Hein^4^

Suzanne M. Herd^1^

Susan S. Hughes^5^

Mohammed Ilyas^3,4^

Jill Jacobson^3,4^

Janda L. Jenkins^5^

Shao Jiang^10^

Jeffrey J. Johnston^1^

Kathryn Keeler^6^

Jonas Korlach^7^

Jennifer Kussmann^5^

Christine Lambert^7^

Caitlin Lawson^5^

Jean-Baptiste Le Pichon^4^

James Steven Leeder^1^

Vicki C. Little^4^

Daniel A. Louiselle^1^

Michael Lypka^10^

Brittany D. McDonald^1^

Neil Miller^1,3,11^

Ann Modrcin^4^

Annapoorna Nair^1^

Shelby H. Neal^1^

Christopher M. Oermann^4^

Donna M. Pacicca^6^

Kailash Pawar^4^

Nyshele L. Posey^1^

Nigel Price^6^

Laura M.B. Puckett^1^

Julio F. Quezada^3,4^

Nikita Raje^3,12^

William J. Rowell^7^

Eric T. Rush^3,5,13^

Venkatesh Sampath^14^

Carol J. Saunders^1,2,3^

Caitlin Schwager^5^

Richard M. Schwend^6^

Elizabeth Shaffer^4^

Craig Smail^1^

Sarah Soden^4^

Meghan E. Strenk^5^

Bonnie R. Sullivan^5^

Brooke R. Sweeney^3,4^

Jade B. Tam-Williams^4^

Adam M. Walter^1^

Holly Welsh^5^

Aaron M. Wenger^7^

Laurel K. Willig^4^

Yun Yan^3,4^

Scott T. Younger^1^

Dihong Zhou^5^

Tricia N. Zion^1,3,4,5^

Isabelle Thiffault^1,2,3^

Tomi Pastinen^1,3^

Affiliations:

1. Genomic Medicine Center, Children’s Mercy Research Institute and Children’s Mercy Kansas City, Kansas City, MO, USA
2. Department of Pathology and Laboratory Medicine, Children’s Mercy Kansas City, Kansas City, MO, USA
3. UKMC School of Medicine, University of Missouri Kansas City, Kansas City, MO, USA
4. Department of Pediatrics, Children’s Mercy Kansas City, Kansas City, MO, USA
5. Division of Genetics, Children’s Mercy Kansas City, Kansas City, MO, USA
6. Department of Orthopedic Surgery, Children’s Mercy Kansas City, Kansas City, MO, USA
7. Pacific Biosciences of California, Inc, Menlo Park, CA, USA
8. PhenoTips, Toronto, Canada
9. Children’s Mercy Research Institute, Kansas City, MO, USA
10. Bionano Genomics, Inc, San Diego, CA, USA
11. Division of Allergy Immunology Pulmonary and Sleep Medicine, Children’s Mercy Kansas City, Kansas City, MO, USA
12. Division of Neonatology, Children’s Mercy Kansas City, Kansas City, MO, USA
13. Department of Internal Medicine, University of Kansas School of Medicine, Kansas City, MO, USA
14. Division of Neonatology, Children's Mercy Hospital Kansas City, Kansas City, MO, USA
